# Adjusting for hidden biases in sexual behaviour data: a mechanistic approach

**DOI:** 10.1101/2023.08.16.23294164

**Authors:** Jesse Knight, Siyi Wang, Sharmistha Mishra

## Abstract

**Background:** Two required inputs to mathematical models of sexually transmitted infections are the average duration in epidemiological risk states (e.g., selling sex) and the average rates of sexual partnership change. These variables are often only available as aggregate estimates from published cross-sectional studies, and may be subject to distributional, sampling, censoring, and measurement biases.

**Methods:** We explore adjustments for these biases using aggregate estimates of duration in sex work and numbers of reported sexual partners from a published 2011 survey of female sex worker in Eswatini. We develop adjustments from first principles, and construct Bayesian hierarchical models to reflect our mechanistic assumptions about the bias-generating processes.

**Results:** We show that different mechanisms of bias for duration in sex work may “cancel out” by acting in opposite directions, but that failure to consider some mechanisms could over- or underestimate duration in sex work by factors approaching 2. We also show that conventional interpretations of sexual partner numbers are biased due to implicit assumptions about partnership duration, but that unbiased estimators of partnership change rate can be defined that explicitly incorporate a given partnership duration. We highlight how the unbiased estimator is most important when the survey recall period and partnership duration are similar in length.

**Conclusions:** While we explore these bias adjustments using a particular dataset, and in the context of deriving inputs for mathematical modelling, we expect that our approach and insights would be applicable to other datasets and motivations for quantifying sexual behaviour data.

## 1 Introduction

Mathematical models of sexually transmitted infections require quantitative estimates of sexual behaviour for model inputs (parameters) [1]. In such models with risk heterogeneity — i.e., considering states that experience differential risks — two important parameters are: the duration of time within an epidemiological risk state/group (or period/season of risk) and the rate of sexual partnership change (often stratified by partnership type) [2, 3, 4, 5]. For example, the average duration of time engaged in sex work can be used to define the modelled rate of “turnover” among sex workers [4]. Similarly, the numbers of main, casual, transactional, and/or paying sexual partners per year can be used to define the modelled rate of infection incidence [6].

Data to inform these parameters largely come from cross-sectional studies, and are often only available as aggregate estimates (vs individual-level data). Such estimates may be subject to several “hidden” sources of bias which can easily go unnoticed, including distributional, sampling, censoring, and measurement biases. Our aim is therefore to explore bias adjuments for estimating: (1) duration in a risk state, and (2) rate of partnership change, from aggregate cross-sectional survey data. We explore and demonstrate these topics using data from a 2011 female sex worker survey in Eswatini [7], to support parameterization of a mathematical model of heterosexual HIV transmission.

## 2 Methods

### Data Source

Full details of the survey methodology are available in [8]. Briefly, 328 women aged 15+ who reported exchanging or selling sex for money, favors, or goods in the past 12 months were recruited via respondent-driven sampling (RDS) [9].

### Approach

We conceptualize bias adjustments to the given data using Bayesian hierarchical models. Specifically, we define explicit distributions for the unbiased data and bias-generating mechanisms, and infer the parameters of these distributions based on the available data, using Gibbs sampling [10]. Implementation details are given in Appendix A.2. Figure 1 gives some supporting diagrams, while Figure 2 illustrates the complete models.

**Figure 1.**
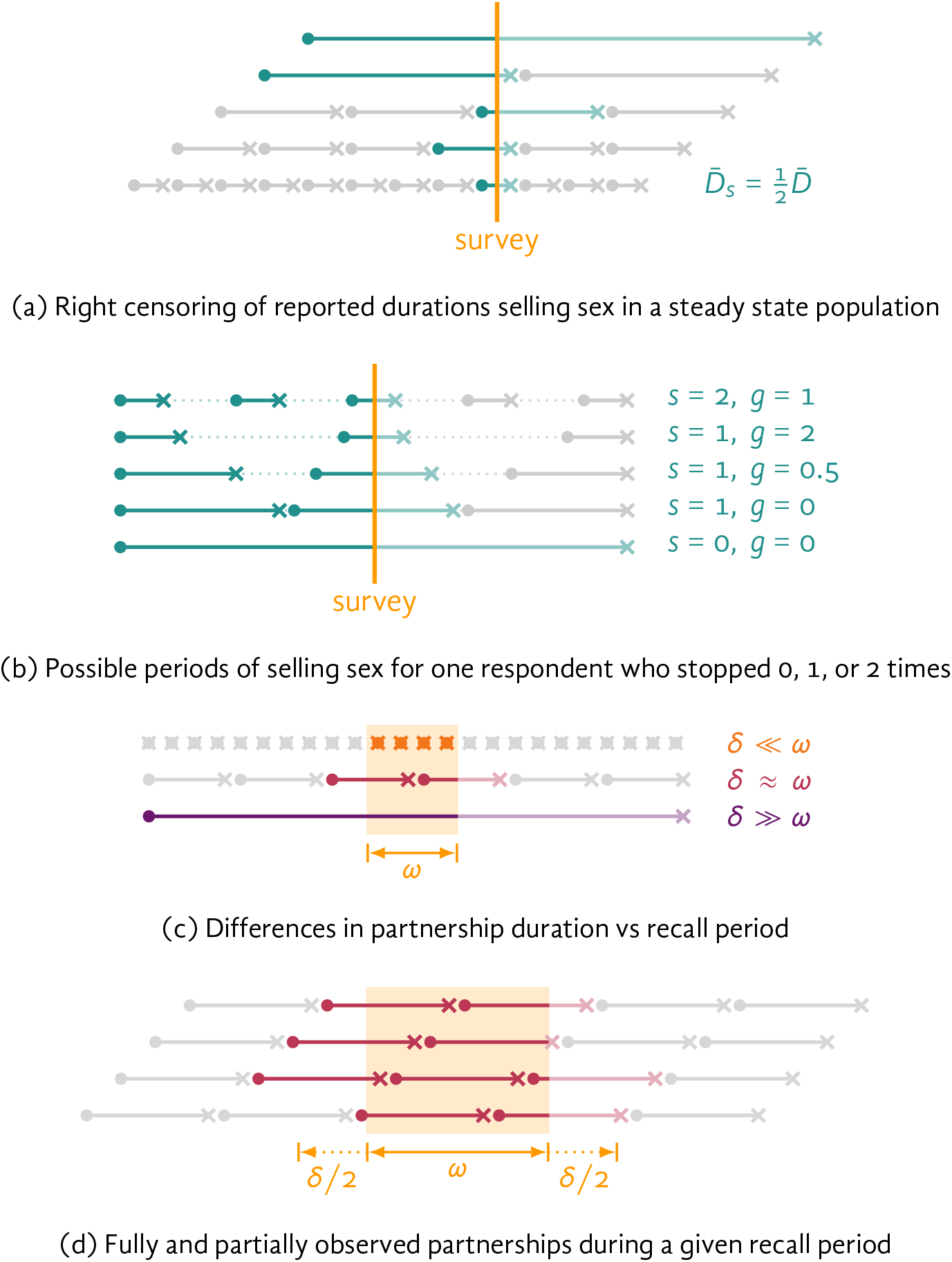
: Diagrams of fully observed, censored, and unobserved periods selling sex or within ongoing sexual partnerships Guide: • : start, × : end, yellow: survey/recall period, full colour: fully observed, faded colour: right censored, grey: unobserved, 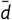 : mean duration at survey and 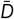 : overall, *s*: number of times stopped selling sex, *g*: relative gap length vs *D, ω*: recall period, *δ*: partnership duration, *x*: number of reported partnerships.

**Figure 2.**
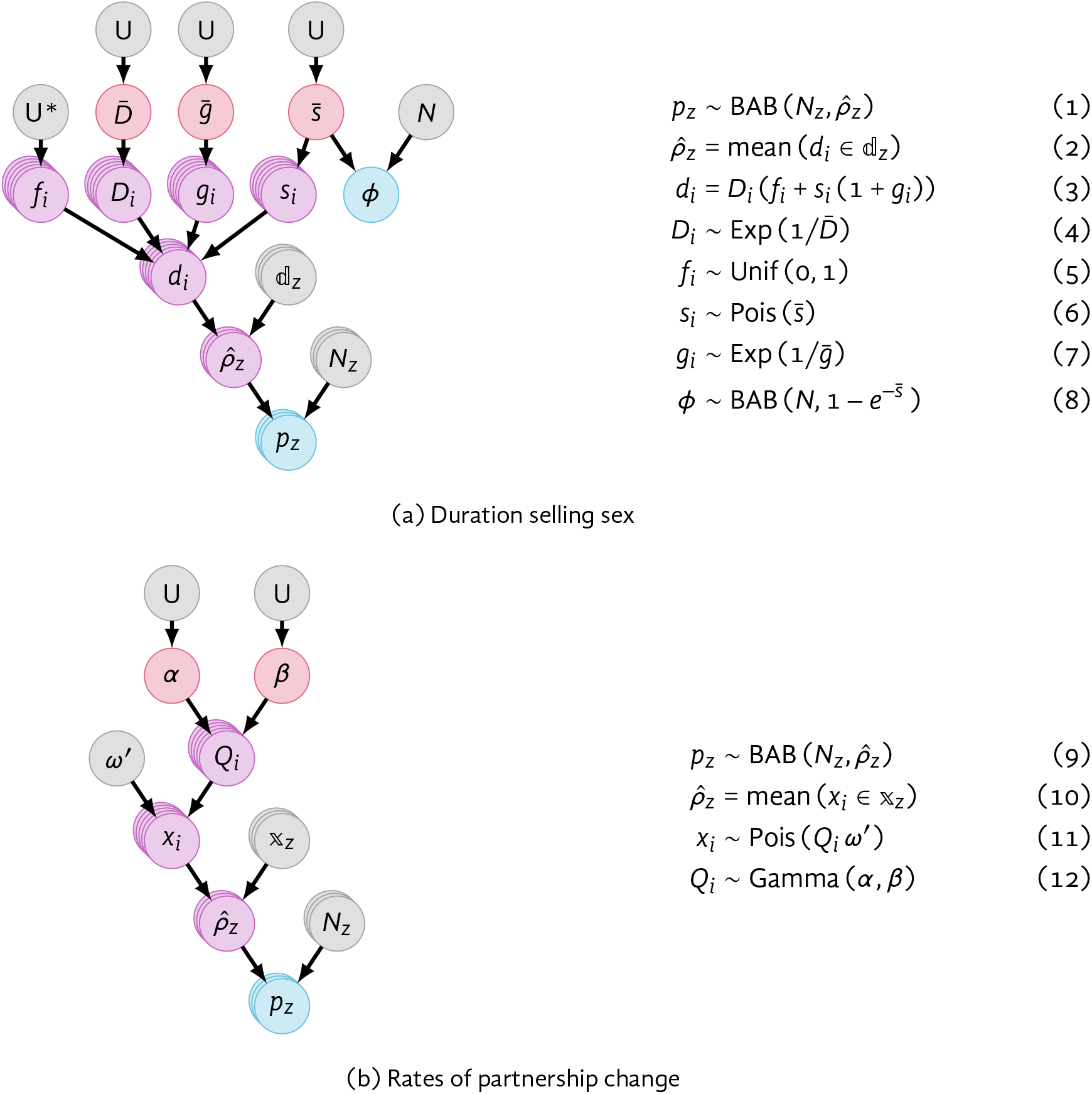
: Graphical and mathematical representations of the proposed Bayesian hierarchical models Guide: gray: fixed variable/distribution, red: target, purple: intermediate, blue: observed. Variables: *p*_*z*_: proportion of population, *N*_*z*_: effective sample size, 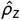: empirically estimated *p*_*z*_ mean, ⅆ_z_: range of reported durations selling sex, *d*_*i*_: reported duration at survey, *D*_*i*_: total (eventual) duration, *f*_*i*_: censoring fraction, *s*_*i*_: number of times stopped selling sex, *g*_*i*_: relative gap length, 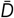: true *D* mean, 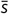: true *s* mean, 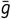 : true *g* mean, *ϕ*: proportion who stopped selling sex at least once, 𝕩: range of reported partner numbers, *x*_*i*_: reported partner numbers, *Q*_*i*_: partnership change rate, *ω*^′^: effective recall period, *α, β* : parameters of *Q*_*i*_ distribution. Distributions: U: uniform / uninformative, BAB: beta approximation of binomial distribution (see § A.3).

### 2.1 Duration Selling Sex

#### Crude Estimates

The survey [7] included questions about the current respondent’s age and the age of first selling sex. The difference between these ages could be used to define a crude “duration selling sex”. Using this approach, the crude median duration was 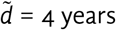 years. However, if durations are assumed to be exponentially distributed — a implicit assumption in compartmental models [11]— then the crude mean could be estimated from the crude median as 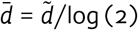 due to skewness. To move beyond crude estimates, next we develop the hierarchical model, considering the following potential biases.

#### Sampling

Sampling bias was considered via RDS-adjustment in [7], yielding mean and 95% CI estimates of the proportions of respondents *p*_*z*_ who had sold sex starting ⅆ_z_ ∈ {0–2, 3–5, 6–10, 11+} years ago (Table A.1, “*z*” enumerates strata). We start by defining a model to identify distributions of reported durations *d*_*i*_ which are consistent with these data. We model each proportion *p*_*z*_ as a random variable with a beta approximation of binomial (BAB) distribution (see Appendix A.3) with parameters *N*_*z*_ and *ρ*_*z*_. We model each *N*_*z*_ as a fixed value, which we fit to the 95% CI of *p*_*z*_ as described in § A.3. We then model each *ρ*_*z*_ as the proportion of reported durations *d*_*i*_ within the interval ⅆ_z_. Since these proportions are difficult to define analytically, we estimate 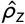 = mean (*d*_*i*_ ∈ ⅆ_z_) from *N* = 100 samples [13].

#### Censoring

These reported durations *d*_*i*_ are effectively right censored because they only capture engagement in sex work up until the survey, and and not additional sex work after the survey (Figure 1a) [12]. If we assume that the survey reaches respondents at a random time point during their total (eventual) duration selling sex *D*_*i*_, we can model this censoring via a random fraction *f*_*i*_ ∼ Unif (0, 1), such that *d*_*i*_ = *f*_*i*_*D*_*i*_; the expected means are then related by 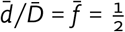.

#### Measurement

Finally, respondents may not sell sex continuously. Reported durations *d*_*i*_ may therefore include multiple periods of selling sex with gaps in between, whereas we aim to model *D*_*i*_ as the durations of individual periods selling sex. Respondents in [7] were not asked whether they ever temporarily stopped selling sex, but a later survey [14] indicated that *ϕ*= 45% had stopped at least once. We model the number of times a respondent may temporarily stop selling sex as a Poisson-distributed random variable *s*_*i*_ with mean 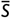. The expected value of *ϕ*given 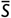is then 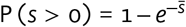. Since *ϕ*= 45% is an imperfect observation, we model *ϕ*as a random variable with a BAB distribution having parameters *N* = 328 and 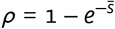, which allows inference on 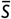 given *ϕ*.

Next, we update the model for reported durations as *d*_*i*_ = *D*_*i*_ (*f*_*i*_ + *s*_*i*_ (1 + *g*_*i*_)), where *g*_*i*_ is the relative duration of gaps between selling sex, with the following rationale. If *s*_*i*_ = 0, then *d*_*i*_ = *f*_*i*_*D*_*i*_ as before, reflecting the censored current period only. If *s*_*i*_ > 0, then *d*_*i*_ also includes *s*_*i*_ prior periods selling sex and the gaps between them (Figure 1b) — i.e., *s*_*i*_ (*D*_*i*_ + *g*_*i*_*D*_*i*_) = *D*_*i*_ *s*_*i*_ (1 + *g*_*i*_). The major assumption we make here is that all successive periods are of equal length, and likewise for gaps between them. We must also assume a distribution for *g*_*i*_, for which we choose 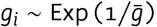, arbitrarily.

#### Summary

Figure 2a summarizes the proposed model graphically. The primary parameter of interest is the mean duration selling sex (for a given period) 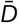, but we must also infer the mean number of times respondents stop selling sex 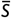, and the mean relative duration of gaps 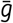. We assume uninformative priors for these 3 parameters.

### 2.2 Rates of Partnership Change

#### Data & Assumptions

The survey [7] also asked respondents to report their numbers of sexual partners (*x*) in a recall period (*ω*) of 30 days. Numbers were stratified by three types of partner: new paying clients, regular paying clients, and non-paying partners. We assume that only a small proportion of new clients go on to become regular clients; thus, we conceptualize “new” clients as effectively “one-off” clients.^1^ Since no survey questions asked about partnership durations (*δ*), we further assume that these were: 1 day with new paying clients, 4 months with regular paying clients, and 3 years with non-paying partners. We now develop the hierarchical model to estimate the expected rate of partnership change for each type, considering the following potential biases.

#### Sampling

As before, [7] estimates RDS-adjusted proportions of respondents *p*_*z*_ (mean, 95% CI) reporting different numbers/ranges of partners 𝕩_*z*_ in the past 30 days (Table A.1). Thus, we take the same approach as in § 2.1 to identify distributions of reported partner numbers *x*_*i*_ which are consistent with the data for each partnership type.

#### Interpretation

Numbers of reported partners (*x*) have generally been interpreted in two ways —*x*/*ω* as the *rate* of partnership change (*Q*) or *x* as the *number* of current partners (*K*):

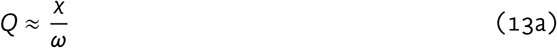

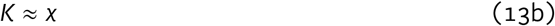

Both interpretations are reasonable under certain conditions: If partnership duration is short and the recall period is long (*δ* ≪ *ω*, e.g., 1 day vs 1 month), then reported partnerships mostly reflect *complete* partnerships, and thus *x*/*ω* ≈ *Q*. If partnership duration is long and the recall period is short (*δ* ≫ *ω*, e.g., 1 year vs 1 month), then reported partnerships mostly reflect *ongoing* partnerships, and thus *x* ≈ *K*. However, if partnership duration and recall period are similar in length (*δ* ≈ *ω*, e.g., 1 month vs 1 month), then reported partnerships reflect a mixture of tail-ends, of complete, and of ongoing partnerships. Thus *x*/*ω* overestimates *Q*, but *x* also overestimates *K*. These three cases are illustrated in Figure 1c.

To adjust for this bias, we again assume that survey/recall period timing is effectively random. Then, if the *end* of the recall period would intersect an ongoing partnership, then a random fraction *f*_*i*_ ∼ Unif (0, 1) of the partnership duration *δ* would be outside the recall period. As before, the expected value 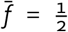. The same goes for the *start* of the recall period. Thus, the recall period is effectively extended by half the partnership duration *δ*/2 on each end, and *δ* overall [15], as illustrated in Figure 1d. We can therefore define unbiased estimators of *Q* and *K* as:

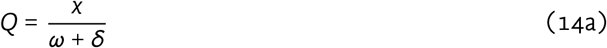

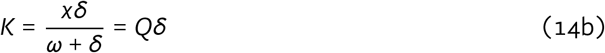

To apply (14) in the hierarchical model, we sample the true rate of partnership change from an assumed distribution *Q*_*i*_ ∼ Gamma (*α, β*), with unknown parameters *α, β*. Then, we model the numbers of reported partners *x*_*i*_ given *Q*_*i*_ and *ω*′ = (*ω* + *δ*) as: *x*_*i*_ ∼ Poi (*Qω*′).

#### Summary

Figure 2b summarizes the proposed model graphically. The primary parameters of interest are *α, β*, which govern the distribution of rates of partnership change (for a given type) *Q*. We assume uninformative priors for these 2 parameters.

#### Comparing Assumptions

To quantify the influence of using the biased vs unbiased estimators of *Q* and *K*, we fit the proposed model for each partnership type under three assumptions: assuming *short* partnerships as in (13a) with *ω*′ = *ω*; assuming *long* partnerships as in (13b) with *ω*′ = *δ*; and *no* assumption on partnership duration as in (14) with *ω*′ = *ω* + *δ*. To illustrate more general trends in the magnitude of bias, we further compared biased vs unbiased estimates of *Q* and *K* across a range of different partnership durations *δ* ∈ [0.1, 10] and recall periods *ω* ∈ [0.1, 10], with fixed true rate *Q* = 1 (arbitrary units).

## 3 Results

### 3.1 Risk Group Duration

Figure A.1 illustrates the distributions of observed proportions *p*_*z*_ vs inferred proportions 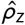 of respondents reporting durations *d*_*i*_ ∈ ⅆ_z_ selling sex, following each stage of adjustment from § 2.1. Figure 3 illustrates the estimated cumulative distributions for years selling sex following each stage of adjustment, while Table A.2 provides the corresponding distribution means 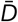 and 95% CI. In this case, the final estimate of 4.06 (2.29, 6.34) is similar to the original median of 4, because each adjustment alternates betwen increasing and decreasing 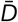. The censoring adjustment yields the largest increase, while the measurement adjustment yields the largest decrease.

**Figure 3.**
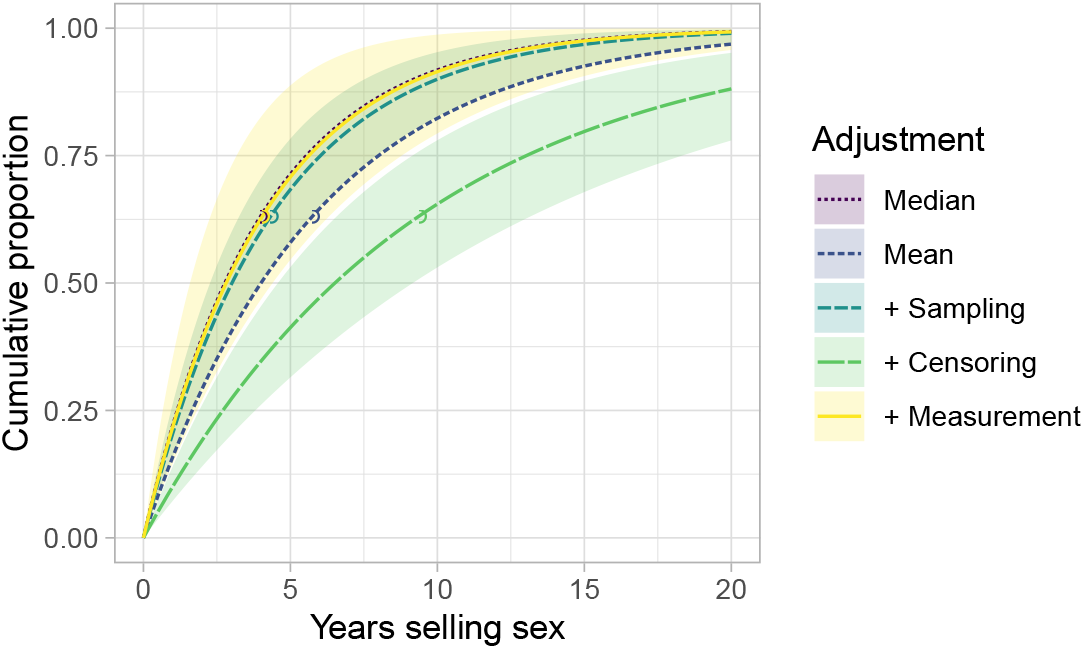
: Estimated cumulative distribution for years selling sex following stages of adjustment Guide: lines: cumulative distribution under posterior mean, shaded ribbon: 95% CI, circles: posterior mean.

### 3.2 Rates of Partnership Change

Figure A.2 illustrates the distributions of observed proportions *p*_*z*_ vs inferred proportions 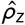 of women reporting *x*_*i*_ ∈ ⅆ_z_ partners in the past 30 days, under the three partnership duration assumptions. Figure 4 illustrates the inferred rates of partnership change (*Q*) and numbers of current partners (*K*) under each assumption, while Table A.3 provides the corresponding means and 95% CI. The biased estimates of *Q* and *K* appear equal because *Q* is defined as per-month. Biases are largest for *Q* with long partnerships (e.g., non-paying partners) and *K* with short partnership (e.g., new clients). However, biases are also large for both *Q* and *K* with “medium-length” partnerships (e.g., regular clients). Figure 5 illustrates generalized trends in these biases.

**Figure 4.**
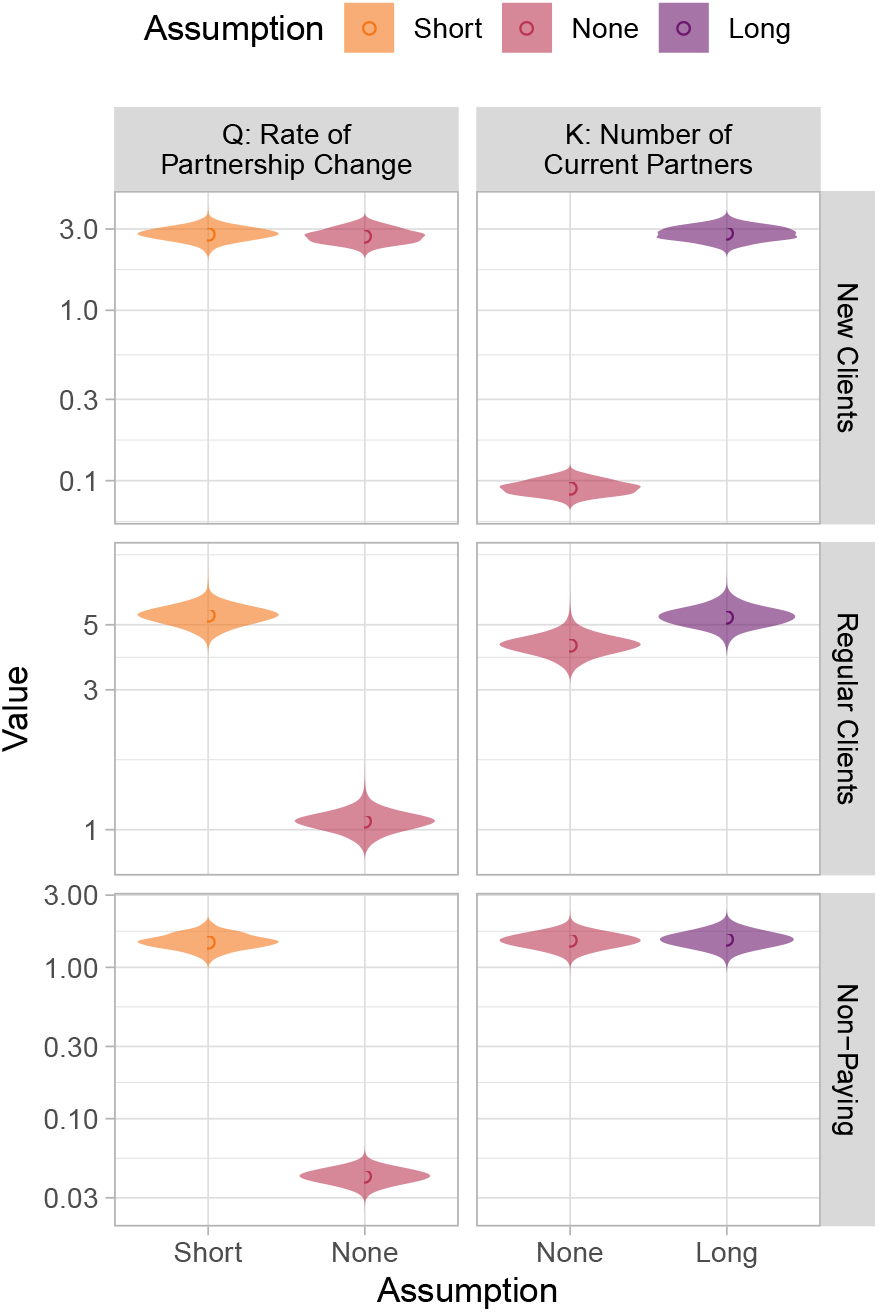
: Estimates of rates of partnership change and numbers of current partners under different partnership duration assumptions for three partnership types reported by female sex workers Guide: circles: posterior mean, shaded area: posterior distribution. Rates are per-month.

**Figure 5.**
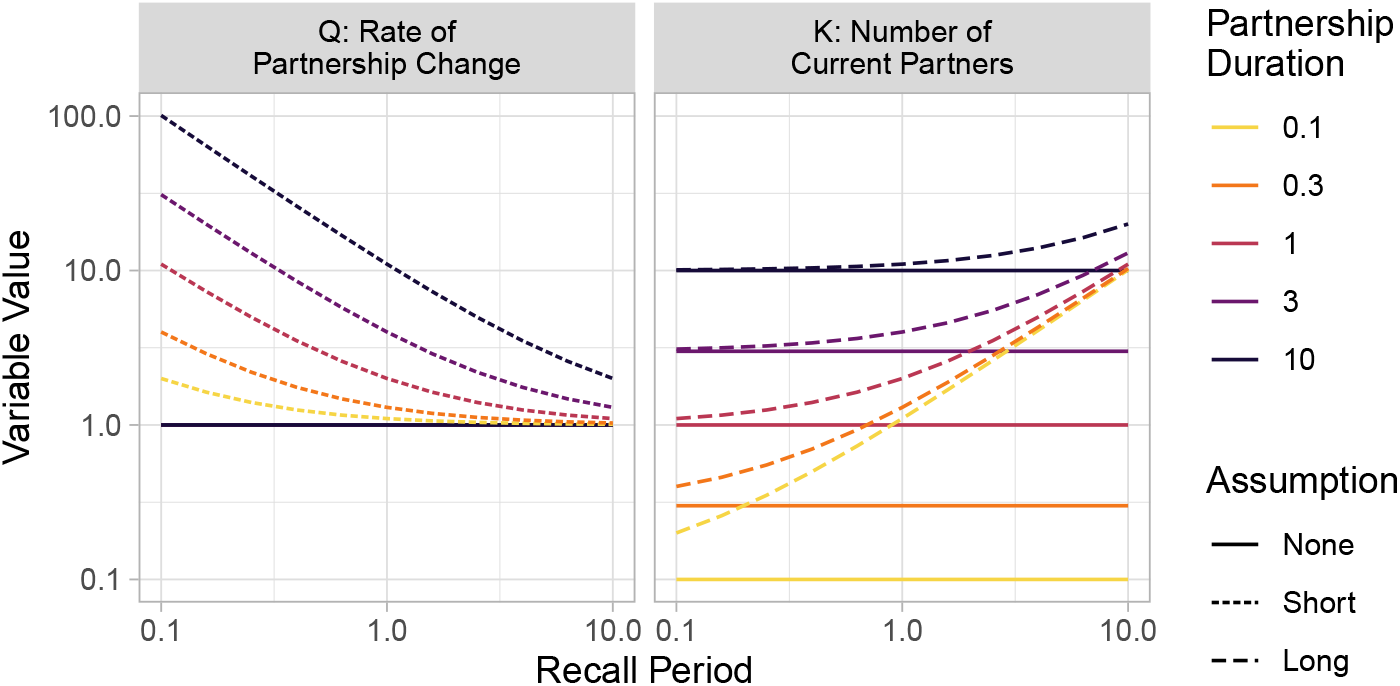
: Estimates of rates of partnership change and numbers of current partners under different partnership duration assumptions for different recall periods and partnership durations Units are arbitrary.

## 4 Discussion

We sought to develop bias adjustments for estimating the mean duration in epidemiological risk states (or periods of risk) and rates of sexual partnership change from aggregate cross-sectional data. We developed these adjustments using Bayesian hierarchical models to incorporate uncertainty in the available data and mechanistic assumptions about several “hidden” bias-generating processes. We showed that these adjustments can influence estimated variable means by factors approaching 2, suggesting that unadjusted estimates of these variables should be interpreted carefully.

We grounded our study in the analysis of aggregate sex work data to parameterize a mathematical model of HIV transmission. However, our approach should be broadly applicable to analysis of other intermittent risk exposures and event rates, including analysis of individual-level data for conventional statistical models. For example, periods of hazardous conditions may need to be quantified in an empiric study of workplace injury risk. Additionally, estimates of population-attributable fractions may be improved through our insight that: in some cross-sectional studies, reported exposure duration reflects only half of the total expected exposure duration.

Our work can also be built upon by considering further potential sources of bias and/or uncertainty. For example, we assumed a fixed duration for each sexual partnership type, but this duration could be modelled as another random variable whose distribution could also be inferred. Moreover, future work could consider rounding error [16], recall bias [17], reporting bias [18], and the like [19].

## Data Availability

All analysis code is available on GitHub, from which all results can be reproduced.

https://github.com/mishra-lab/hidden-bias-sex-data

## Acknowledgements

We thank: Linwei Wang, Korryn Bodner, Amrita Rao, Kate Rucinski, Le Bao, and Stefan Baral for helpful discussions on epidemiological implications of the work; Jarle Tufto and Michael Neely for guidance on mathematical modelling of censored durations and Poisson processes.

## Footnotes

1 The number of new clients per recall period could also be used to define a rate of partnership change [12], but we do not explore this approach here.

