## Appendix for "Adjusting for hidden biases in sexual behaviour data: a mechanistic approach"

**Date** August 16, 2023

### A Supplement

#### A.1 Source Data

Table A.1 gives the RDS-adjusted data from [1].

Table A.1: RDS-adjusted proportions for variables of interest

| Variable | Stratum | Mean | (95% CI) |
|---|---|---|---|
| Years selling sex | 0–2 | 38.3 | (27.5, 49.1) |
| | 3–5 | 32.1 | (23.6, 40.7) |
| | 6–10 | 20.2 | (13.2, 27.1) |
| | 11+ | 9.4 | (04.4, 14.4) |
| New clients [a] | 0–1 | 16.4 | (09.8, 23.0) |
| | 2 | 43.4 | (33.3, 53.5) |
| | 3 | 15.2 | (09.6, 20.9) |
| | 4 | 13.1 | (07.0, 19.2) |
| | 5 | 11.8 | (06.0, 17.6) |
| Regular clients [a] | 0–1 | 10.0 | (01.9, 18.1) |
| | 2 | 8.5 | (03.2, 13.8) |
| | 3 | 15.9 | (09.8, 21.9) |
| | 4 | 10.0 | (04.5, 15.6) |
| | 5 | 8.1 | (03.8, 12.3) |
| | 6 | 10.7 | (05.8, 15.5) |
| | 7+ | 36.9 | (26.4, 47.3) |
| Non-paying partners [a] | 0 | 12.5 | (04.8, 20.1) |
| | 1 | 50.8 | (42.9, 58.7) |
| | 2 | 23.6 | (16.8, 30.3) |
| | 3+ | 13.2 | (07.2, 19.1) |

[a] Number reported in the past 30 days. Data from [1].

#### A.2 Code

All analysis code is available online at: `github.com/mishra-lab/hidden-bias-sex-data`.
We fit the Bayesian hierarchical models using rjags: `cran.r-project.org/package=rjags`, with 1000 adaptive iterations and 100,000 sampling iterations.

#### A.3 Beta Approximation of the Binomial Distribution

The distributions of RDS-adjusted variables in [1] were reported as adjusted proportions (mean, 95% CI) for different stratifications of the variable value; e.g., 16.4 (9.8, 23.0) % of respondents reported 0–1 new clients in the past 30 days. For each proportion, we defined a beta approximation of the binomial (BAB) distribution:

$$P(\rho) = \frac{\Gamma(\alpha+\beta)}{\Gamma(\alpha)\Gamma(\beta)} \rho^{\alpha-1}(1-\rho)^{\beta-1} \approx \binom{N}{n} \rho^{n}(1-\rho)^{N-n} \tag{A.1}$$

with $\alpha = N\rho$ and $\beta = N(1-\rho)$. We fixed $\rho$ as the adjusted point estimate, and estimated $N$ by minimizing the sum of squared differences between the 95% quantiles of (A.1) given $N$ and the reported 95% CI for the adjusted proportion.

#### A.4 Risk Group Duration

**Fitting to RDS-Adjusted Proportions.** Figure A.1 illustrates the observed vs inferred proportions of respondents reporting different durations selling sex, following each stage of adjustment from § 2.1.

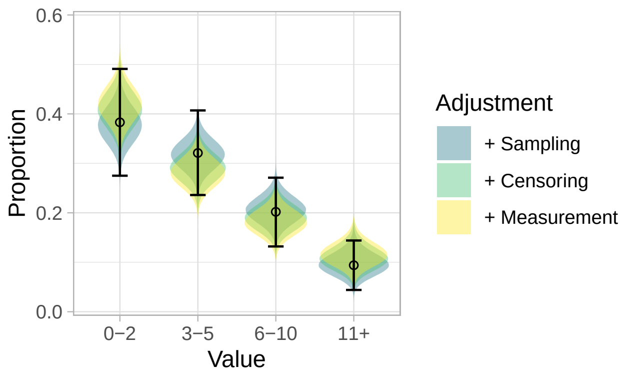


Figure A.1: Proportions of respondents reporting different durations selling sex: observed (points and ranges) vs inferred posterior (coloured regions) after 3 stages of adjustment

**Numeric Summary.** Table A.2 summarizes the estimated exponential distribution means (95% CI) for years selling sex following each stage of adjustment from § 2.1.

Table A.2: Estimated mean durations selling sex (years) following each stage of adjustment

| Adjustment | Mean | (95% CI) |
|---|---|---|
| Median | 4.00 | — |
| Mean | 5.77 | — |
| + Sampling | 4.35 | (3.27, 5.72) |
| + Censoring | 9.40 | (6.60, 13.22) |
| + Measurement | 4.06 | (2.29, 6.34) |

#### A.5 Rate of Partnership Change

**Fitting to RDS-Adjusted Proportions.** Figure A.2 illustrates the observed vs inferred proportions of respondents reporting different numbers of partners in the past 30 days, under each partnership duration assumption from § 2.2.

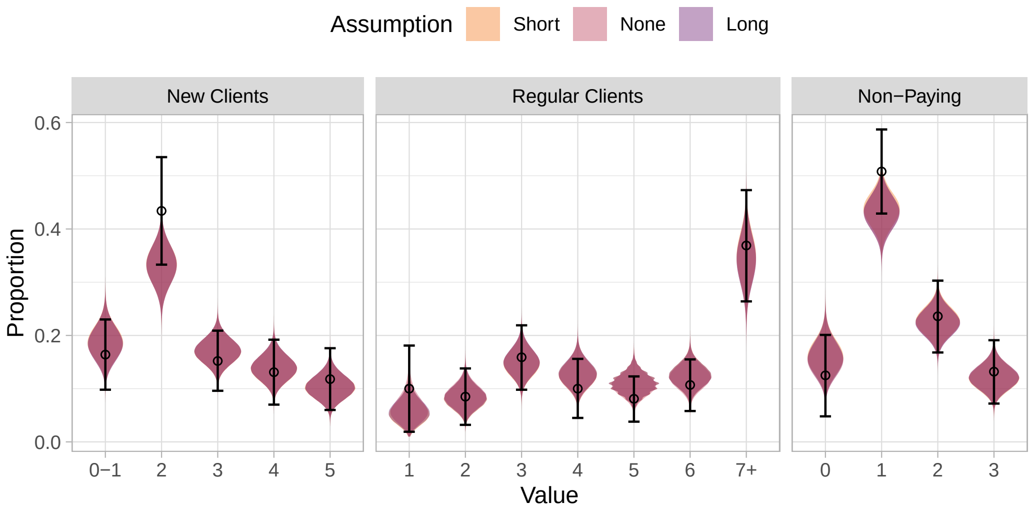


Figure A.2: Proportions of respondents reporting different numbers of partner in the past 30 days: observed (points and ranges) vs inferred posterior (coloured regions) under different partnership duration assumptions

**Numeric Summary.** Table A.3 summarizes the means (95% CI) for rates of partnership change and numbers of current partners estimated under each partnership duration assumption from § 2.2.

Table A.3: Biased vs unbiased estimates of rates of partnership change and numbers of current partners for three partnership types

| | | Rate $Q$ [a] | | Number $K$ | |
|---|---|---|---|---|---|
| Partnership Type | Bias [b] | Mean | (95% CI) | Mean | (95% CI) |
| New Clients | Biased | 2.82 | (2.33, 3.35) | 2.84 | (2.36, 3.37) |
| | Unbiased | 2.75 | (2.29, 3.31) | 0.09 | (0.08, 0.11) |
| Regular Clients | Biased | 5.38 | (4.60, 6.20) | 5.33 | (4.57, 6.19) |
| | Unbiased | 1.07 | (0.90, 1.25) | 4.28 | (3.62, 5.02) |
| Non-Paying | Biased | 1.49 | (1.17, 1.86) | 1.54 | (1.20, 1.95) |
| | Unbiased | 0.04 | (0.03, 0.05) | 1.51 | (1.18, 1.88) |

[a] Rates are per-month; [b] biased $Q$ assume short partnerships as in (13a); biased $K$ assume long partnerships as in (13b).
